# Machine Learning Analysis of PD-L1 and CD8 Expression Patterns in Penile Squamous Cell Carcinoma: Implications for Immunotherapy

**DOI:** 10.1101/2024.12.11.24318853

**Authors:** Sofia Canete-Portillo, Diana Taheri, María del Carmen Rodriguez-Peña, Marie-Lisa Eich, Margaret Cocks, Antonio L. Cubilla, George J. Netto, Alcides Chaux

## Abstract

**Introduction:** In 2023, the US reported an estimated 2,050 new penile squamous cell carcinoma (SCC) cases, with mortality rates rising to 23%. Despite therapeutic advancements, the need to identify molecular and immunotherapeutic targets, especially PD-L1 and CD8, has become increasingly evident as immunotherapy emerges as a promising treatment strategy.

**Methods:** Tissue samples from 108 SCC patients were examined using four tissue microarrays (TMA). Alongside the assessment of histologic features and PD-L1 and CD8 expressions, a machine learning model was employed. This model, constructed in Python, leveraged various statistical tests and methodologies to discern the predictive relationships between PD-L1/CD8 expression and pathologic features, with potential implications for immunotherapy selection.

**Results:** Histologic subtype was predominant in PD-L1 expression in tumor cells, explaining 13.5% variability, whereas combined with host response, it reached 20%. For intratumoral lymphocytes, host response stood out, explaining 11.2% of PD-L1 and 15% of CD8 variability. In peritumoral lymphocytes, host response again dominated, accounting for 21.1% of CD8 variability. These findings align with recent studies showing the importance of immune markers in predicting treatment response and prognosis.

**Conclusions:** The study highlighted the crucial relationship between penile SCC’s pathologic features and PD-L1/CD8 expression, emphasizing their potential as both prognostic and predictive biomarkers. By integrating traditional histopathology with machine learning, we provide valuable insights for patient stratification and personalized immunotherapeutic strategies in penile SCC.

## INTRODUCTION

In 2023, the US reported an estimated 2,050 new cases of penile squamous cell carcinoma (SCC), a rare tumor (Siegel et al., 2023). The percentage of deaths due to this condition rose from 16% in 2016 to 23% in 2023 (Siegel et al., 2016, 2023). While there are limited effective treatment options, the standard treatment can significantly affect psychological, urinary, and sexual functions (Maddineni et al., 2009; Zukiwskyj et al., 2013). Organ-sparing techniques and other treatments have been introduced for specific patients (Tang et al., 2018). Nevertheless, for those with advanced disease, the prevailing treatments are partial or total penectomy, often combined with systemic chemotherapy and radiotherapy (Marchioni et al., 2018). Efforts are underway to find alternatives that reduce the psychological and anatomical consequences of partial/total penectomies, which are currently the mainstay for advanced cases. Consequently, there is a pressing need to identify new molecular and immunotherapeutic targets (Giunchi et al., 2021; Vanthoor et al., 2021).

Programmed death-ligand 1 (PD-L1) plays a pivotal role in immune system regulation. It is a coinhibitory molecule that, when engaged, can dampen the T-cell response. This is achieved by suppressing T-cell proliferation and reducing cytokine production, which are crucial for a robust immune response. Interestingly, many tumor cells have developed a cunning mechanism to exploit this system (Gou et al., 2020). By up-regulating PD-L1, these tumor cells can effectively cloak themselves, evading detection and attack by the host immune system (Liu et al., 2019; Rosenbaum et al., 2016).

Recently, the advent of immune-checkpoint inhibitors has revolutionized cancer therapy. These drugs have shown remarkable efficacy against various tumor types by essentially “unmasking” the tumors to the immune system. In penile SCC, understanding of PD-L1 expression has advanced significantly, with studies reporting positivity rates ranging from 40% to 62% (Su et al., 2020; Warli et al., 2022), indicating substantial potential for PD-1/PD-L1-targeted therapies. This variability in expression underscores the importance of routine PD-L1 testing in clinical settings to identify candidates for immunotherapy effectively.

Recent research has also delved into the role of CD8+ T cells in various cancers, highlighting their potential significance in tumor prognosis and therapeutic strategies (Ao et al., 2023; R. Li et al., 2020; Y. Li et al., 2019). While specific studies on CD8 expression in penile cancer are limited, the broader context of CD8 role in oncology offers insights. Higher densities of CD8+ TILs correlate with improved overall survival and disease-free survival in various cancers, including penile cancer (Ottenhof et al., 2018; Vries et al., 2023). The spatial distribution of CD8+ TILs within the tumor microenvironment has been shown to have prognostic implications, suggesting that not only the quantity but also the localization of CD8+ TILs could influence patient prognosis.

The relationship between CD8+ TILs and immunotherapy response has gained particular attention. Studies have demonstrated that patients with higher CD8+ TIL densities may be more likely to benefit from immunotherapeutic approaches (F. Li et al., 2021), a finding that warrants exploration in the context of penile cancer as immunotherapy becomes a more prominent treatment modality. Furthermore, the balance between CD8+ T cells and regulatory T cells (Tregs) in the tumor microenvironment has emerged as a critical determinant of immune response and potential therapeutic success.

In our current research, we aim to bridge this knowledge gap. We have undertaken a comprehensive evaluation of PD-L1 and CD8 expression across a substantial dataset of penile SCC patients. These patients hail from regions where the incidence of the disease is notably high (Chaux et al., 2013; de Sousa et al., 2015). Our primary objective is to discern the relationship between pathologic features and PD-L1 and CD8 expression, to shed light on potential therapeutic avenues and better understand the disease’s immunological landscape. This understanding is particularly crucial as recent studies have highlighted the complex interplay between tumor characteristics, immune markers, and treatment responses, suggesting the potential for more personalized therapeutic approaches based on individual tumor immune profiles.

## MATERIALS AND METHODS

The current study was approved by the Institutional Review Board at the Johns Hopkins School of Medicine (Baltimore, MD).

### Case Selection and Tissue Microarray Construction

The present study includes tissue samples from 108 patients with invasive squamous cell carcinoma of the penis from the consultation files of one of the authors (ALC). Cases were selected based on availability of formalin-fixed, paraffin-embedded tissue blocks. From each case, 1–4 blocks were selected. Four tissue microarrays (TMA) were built at the Johns Hopkins TMA Lab Core (Baltimore, MD) using a previously described procedure (Fedor & De Marzo, 2005). Three tissue cores of 1 mm each were obtained per block, giving a representation of 3–12 spots per case. Normal tissue from various anatomical sites were included as control tissue. A total of 528 TMA spots were evaluated from the 108 cases.

### Morphological Evaluation

Pathologic features were evaluated using H&E-stained tissue sections by three expert genitourinary pathologists. The following pathologic features were evaluated:

#### 1. Histologic subtype

Histologic subtyping was carried out in whole tissue sections using the 5th edition (2022) WHO criteria for classification of tumors of the urinary system and male genital organs (Moch et al., 2022).

#### 2. Histologic grade

Histologic grading was carried out spot by spot using previously published and validated criteria (Chaux, 2015). Briefly, grade 1 tumors were composed of well-differentiated cells, almost undistinguishable from normal squamous cells except for the present of minimal basal/parabasal cell atypia. Grade 3 tumors were composed of any proportion of anaplastic cells showing nuclear pleomorphism, coarse chromatin, prominent nucleolus, irregular and thickened nuclear membrane, abundant and atypical mitoses. Grade 2 tumors corresponded to those cases not fitting criteria for grade 1 or grade 3 (i.e., it was an exclusion category).

#### 3. Host response

Host response was evaluated spot by spot, using previously published criteria (Cocks et al., 2017). Depending on the intensity of the inflammatory infiltrate observed, each spot was classified as showing no inflammation, mild inflammation, moderate inflammation, or intense inflammation.

### Immunohistochemistry

Antibodies were acquired from commercial sources and applied as detailed below. PD-L1 (Cell Signaling, Boston, MA; E1L3N) 1:100 diluted in antibody dilution buffer with a 45-minute incubation at room temperature. CD8 (Thermo Scientific, Waltham, MA; RB-9009-P0) 1:800 diluted in antibody dilution buffer with a 45-minute incubation at room temperature. For CD8, antigen retrieval was done with EDTA buffer (pH 9.0) before antibody incubation. Before incubation with primary antibody, a heat-induced antigen retrieval step was performed for 20 minutes in a steamer with citrate buffer solution (pH 6.0). Detection of immunolabeling was performed using antimouse or antirabbit horseradish peroxidase–conjugated secondary antibodies and counterstaining was performed with 3,3′-diaminobenzidine. All markers were separately assessed in tumor cells as well as associated immune cells.

PD-L1 expression was evaluated in tumor cells and in intratumoral lymphocytes. PD-L1 positivity was defined as the presence of any extent of tumor cell (or immune cell) membranous positivity in one or more representative TMA spots (>0%). Tumor infiltrating lymphocytes were reported as the number of CD8+ lymphocytes in the area of highest density (hotspot) per high-power field (×40) both in tumor (intratumoral lymphocytes) and in stroma (peritumoral lymphocytes).

### Data Analysis

Data was analyzed using Python 3.12.8. Contingency tables were evaluated using the Pearson’s chi-square test. Correlations between numeric variables were evaluated using the Spearman’s rho coefficient. Numeric values were compared in groups using the Kruskal-Wallis test. A 2-tailed P<0.01 was required for statistical significance. These statistical tests were implemented using the SciPy library (Virtanen et al., 2020).

A machine learning model was constructed and validated to investigate the predictive relationship between PDL-1 and CD8 expression and pathologic features (histologic subtype, histologic grade, and inflammatory host response). The data was systematically prepared, with categorical features encoded for numerical compatibility, and subsequently divided into training and testing sets (80% and 20%, respectively). Linear regression was employed to predict pathologic outcomes. The model underwent rigorous training on the training data and was evaluated on the testing data using the explained variance score, a critical metric for assessing predictive performance. We interpreted this metric as the percentage of the variability of the considered marker accounted for the evaluated pathologic features. Machine learning models were built using the Scikit-learn library (Pedregosa et al., 2011).

In adherence to the principles of open science, we have taken measures to ensure full transparency and accessibility of our research analysis. We have made the entire analytic data flow, including the comprehensive Jupyter Notebook containing the Python code, the dataset used, and accompanying scripts, openly accessible to the scientific community. Interested researchers and collaborators can access this resource through the following link: https://github.com/alcideschaux/PFCK-Penis-PRY/.

## RESULTS

### Pathologic Features

The most common subtype, as expected, was usual squamous cell carcinoma (45 cases), followed by warty-basaloid (24 cases), warty (16 cases) and basaloid (11 cases) carcinomas. Other subtypes included papillary (9 cases), verrucous (2 cases) and sarcomatoid (1 case) carcinomas. Grade 1 was observed in 51 spots, grade 2 was observed in 191 spots, and grade 3 was observed in 262 spots. This over-representation of grade 3 tumors is expected, considering the geographical location of the patients. Tumors in patients from geographical areas of high incidence of penile cancer tend to be larger and of higher grade. In most cases, a host response was observed. In only 4 spots, no inflammatory cells were seen. In the remaining cases, mild inflammation was seen in 96 spots, moderate inflammation in 154 spots, and intense inflammation in 250 spots.

Basaloid and sarcomatoid carcinomas were entirely composed of grade 3 areas. Warty-basaloid and warty carcinomas were composed of predominantly grade 2 and grade 3 areas, while papillary and verrucous carcinoma were composed predominantly of grade 1 and grade 2 areas. Usual squamous cell carcinoma showed the heterogeneous aspect that it most common, with a mixture of histologic grades, predominantly grade 2 areas. This distribution pattern was consistent with the typical morphology of penile squamous cell carcinomas regarding histologic subtypes and grades. The association between histologic grade and histologic subtype was statistically significant (P<0.00001).

Intense inflammation predominated across histologic subtypes, showing a similar pattern, with no significant differences between host response and histologic subtypes (P=0.24). Intense inflammation predominated in grade 2 and grade 3 tumors, followed by moderate inflammation and mild inflammation. In grade 1 tumors, proportions of mild, moderate, and intense inflammation were similar. These differences were not statistically significant (P=0.22), indicating no association between histologic grade and host response.

### PD-L1 Expression

#### Overall Expression

PD-L1 expression in tumor cells was evaluable in 504 spots. In tumor cells, mean expression was 26%, with a standard deviation of 34%. The median expression was 5%, with an interquartile range of 40%. The minimum value was 0% and the maximum value was 100%. PD-L1 expression in tumor cells showed a marked right-skewed distribution, indicating that most values were extremely low. Considering > 0% as the threshold for PD-L1 positivity, most spots (63%) were positive, compared to negative spots (37%). This positivity rate aligns with recent studies reporting PD-L1 expression ranges between 40% and 62% in penile SCC (Su et al., 2020; Warli et al., 2022). Two patterns of PD-L1 expression were observed in tumor cells. The predominant pattern was cytoplasmic and membranous (76%) with only cytoplasmic expression in the remaining cases (24%).

PD-L1 expression in intratumoral lymphocytes was evaluable in 497 spots. The minimum and maximum number of PD-L1+ lymphocytes were 0 and 70, respectively. Mean PD-L1 positivity was 7, with a standard deviation of 10 lymphocytes. The median number of positive PD-L1 intratumoral lymphocytes was 5, with an interquartile range of 9 lymphocytes.

A scatterplot of PD-L1 expression in tumor cells vs. intratumoral lymphocytes showed an apparent positive association (see online repository). This positive association was confirmed using Spearman’s correlation test, which showed a statistically significant, moderate positive correlation (rho=0.47, P<0.0001).

#### Expression by Pathologic Features

##### Histologic subtype

When considering the percentage of positive cells, higher expression of PD-L1 in tumor cells were noted for the sarcomatoid, basaloid and warty-basaloid subtypes (median of 100%, 25% and 20%, respectively). Low expression levels were noted in the usual and warty subtypes (5%), while the median expression was 0% for the papillary and verrucous subtypes. These differences were statistically significant (P<0.0001). Regarding the association of histologic subtypes and PD-L1 expression in intratumoral lymphocytes, the median count of PD-L1+ lymphocytes was high in sarcomatoid carcinomas (15 PD-L1+ lymphocytes). It showed moderate values in usual, basaloid, warty, and warty-basaloid carcinomas (5 PD-L1+ lymphocytes), and was low in papillary and verrucous carcinomas (2 PD-L1+ lymphocytes and no PD-L1+ lymphocytes). These differences were also statistically significant (P<0.0001).

##### Histologic grade

Percentages of positive PD-L1 tumor cells increased from grade 1 to grade 2 to grade 3 tumors (median of 0%, 1% and 15%), suggesting an association between PD-L1 positivity in tumor cells and histologic grade. The Kruskal-Wallis test yielded a P<0.0001, indicating that the percentage differences were unlikely to be seen by chance alone. This finding aligns with recent studies showing a correlation between PD-L1 expression and tumor aggressiveness (Sangkhamanon, 2023). A similar trend was observed between histologic grade and PD-L1 expression in intratumoral lymphocytes (median numbers of PD-L1+ lymphocytes of 2, 2 and 5), but it did not reach the threshold for statistical significance (P=0.014).

##### Host response

Median percentage of PD-L1 positive tumor cells increased from mild to intense inflammation. Median expression of PD-L1 in tumor cells was 0% when no inflammation was seen, 1% with mild inflammation, 2% with moderate inflammation and 15% with intense inflammation. This association was statistically significant (P<0.0001). The medium number of PD-L1 positive intratumoral lymphocytes increased from mild to intense inflammation. No PD-L1 positive intratumoral lymphocytes were identified when no inflammation was seen. The median number of PD-L1 positive intratumoral lymphocytes was 1 PD-L1+ lymphocyte with mild inflammation, 2 PD-L1+ lymphocytes with moderate inflammation and 5 PD-L1+ lymphocytes with intense inflammation. This association was statistically significant (P<0.0001).

#### Impact of Pathologic Features

For PD-L1 in tumor cells, histologic subtype explained 13.5% of PD-L1 variability, while histologic grade and host response explained 3.5% and 6.5% of the variability. When 2 features were combined, the highest explanatory power was for histologic subtype + host response (20%), followed by host response + histologic grade (9.6%) and histologic subtype + histologic grade (9.0%). When all 3 features were considered, they explained 16.3% of the PD-L1 variability in tumor cells. Overall, these results suggest that histologic subtype has the highest explanatory power for PD-L1 expression in tumor cells, while histologic grade had the lowest, either by themselves or in combination.

In intratumoral lymphocytes, the scenario was different. First, when considered separately, the highest explanatory power was given by host response (11.2%), followed by histologic subtype (4.1%) and histologic grade (2.5%). When combining 2 features, the highest values were for host response + histologic grade (14.2%) and host response + histologic subtype (13.9%). Histologic subtype + histologic grade explained only 5.6% of the variability. Finally, when the 3 features were combined, the explanatory power was 16.3%, not a significant improvement. Thus, host response was the feature that explained better the variability in PD-L1 expression observed in intratumoral lymphocytes.

### CD8 Expression

#### Overall Expression

CD8 expression in intratumoral lymphocytes was evaluable in 506 spots. In intratumoral lymphocytes, the mean expression was 10 CD8+ lymphocytes, with a standard deviation of 16 CD8+ lymphocytes. The median expression was 4 CD8+ lymphocytes, with an interquartile range of 12 CD8+ lymphocytes. The number of positive lymphocytes ranged from 0 to 120, showing a marked right skewed distribution.

CD8 expression in peritumoral lymphocytes was evaluable in 503 spots. In peritumoral lymphocytes, the mean expression was 27 CD8+ lymphocytes, with a standard deviation of 27 CD8+ lymphocytes. The median expression was 19 CD8+ lymphocytes, with an interquartile range of 35 CD8+ lymphocytes. The number of positive lymphocytes ranged from 0 to 150, showing a right skewed distribution.

A scatterplot of CD8 expression in intratumoral lymphocytes vs. peritumoral lymphocytes showed a positive association (see online repository). This positive association was confirmed using Spearman’s correlation test, which showed a statistically significant, moderate positive correlation (rho=0.37, P<0.0001). This finding aligns with recent studies highlighting the importance of both intratumoral and peritumoral CD8+ T cell infiltration in cancer immunology (Ao et al., 2023).

#### Expression by Pathologic Features

##### Histologic subtype

When considering intratumoral lymphocytes, median positivity was higher in usual, basaloid, warty-basaloid, and sarcomatoid subtypes (6 CD8+ lymphocytes). Lower median expression was observed in papillary (4 CD8+ lymphocytes) and warty (2 CD8+ lymphocytes) subtypes, while no positive lymphocytes were observed in verrucous carcinoma. These differences were statistically significant (P=0.0008). For peritumoral lymphocytes, the median expression was high in most subtypes, including verrucous (23 CD8+ lymphocytes), usual carcinomas (22 CD8+ lymphocytes), warty-basaloid and papillary carcinomas (20 CD8+ lymphocytes), and basaloid carcinomas (19 CD8+ lymphocytes). The median expression was lower in warty and sarcomatoid carcinomas (14 CD8+ lymphocytes). These differences were not statistically significant (P=0.28).

##### Histologic grade

The median number of intratumoral CD8+ lymphocytes increased from grade 1 (1 CD8+ lymphocyte), to grade 2 (2 CD8+ lymphocytes), to grade 3 (6 CD8+ lymphocytes). These differences were statistically significant (P < 0.00001). This association between histologic grade and CD8 expression was not observed in peritumoral lymphocytes. In peritumoral lymphocytes, the median expression was 24 CD8+ lymphocytes in grade 1, 16 CD8+ lymphocytes in grade 2 tumors, and 19 CD8+ lymphocytes in grade 3 tumors. Differences were not statistically significant (P=0.92).

##### Host response

The median number of intratumoral CD8+ lymphocytes increased depending on the intensity of the host inflammatory response. The median number of CD8+ intratumoral lymphocytes was 2 when the host response was mild or moderate, and increased to 8 CD8+ lymphocytes when the response was intense. These differences were statistically significant (P<0.00001). A similar association was observed between median expression of peritumoral CD8+ lymphocytes and host response. The median number of CD8+ peritumoral lymphocytes was 4 CD8+ in mild host response, 12 CD8+ lymphocytes in moderate host response, and 32 CD8+ lymphocytes in intense host response. These differences were statistically significant (P<0.00001). These findings support recent studies showing the importance of host immune response in determining CD8+ T cell infiltration patterns (F. Li et al., 2021).

#### Impact of Pathologic Features

For CD8 in intratumoral lymphocytes, host response explained 15.0% of CD8 variability, while histologic subtype and histologic grade explained 7.7% and 4.7% of CD8 variability. When 2 features were combined, the highest explanatory power was for host response + histologic subtype (19.4%), followed by host response + histologic grade (11.1%) and histologic subtype + histologic grade (5.3%). When all 3 features were combined, they explained 18.7% of CD8 variability. These results suggest that host response has the highest explanatory power for variability in CD8 expression in intratumoral lymphocytes, aligning with recent findings about the importance of the tumor immune microenvironment (Ottenhof et al., 2018).

For CD8 in peritumoral lymphocytes, host response explained 21.1% of CD8 variability, while histologic subtype and histologic grade explained 0.4% and 0.1% of CD8 variability. When 2 features were combined, the highest explanatory power was for host response + histologic grade (23.0%), followed by host response + histologic subtype (20.4%) and histologic subtype + histologic grade (0.01%). When all 3 features were combined, they explained 21.2% of CD8 variability. These results suggest that host response has the highest explanatory power for variability in CD8 expression in peritumoral lymphocytes, a finding that has important implications for immunotherapy response prediction (Vries et al., 2023).

## DISCUSSION

This comprehensive study on PD-L1 and CD8 expression in penile squamous cell carcinoma (PSCC) provides crucial insights into the intricate relationship between pathologic features and immune markers. By employing both traditional histopathological methods and cutting-edge machine learning techniques, our research not only confirms previous findings, but also uncovers novel associations that have significant implications for prognosis and potential therapeutic strategies in PSCC.

Our analysis reveals a complex pattern of PD-L1 expression in both tumor cells and intratumoral lymphocytes, strongly associated with specific histologic subtypes, higher grades, and increased inflammatory host response. These findings build upon our previous observations (Cocks et al., 2017) and align with the broader literature on PD-L1 expression in PSCC. The wide range of PD-L1 positivity rates reported in recent studies, from 40% to 62% (Sangkhamanon, 2023; Su et al., 2020), underscores the critical need for standardized evaluation techniques in clinical practice.

The association between PD-L1 expression and more aggressive disease characteristics reinforces its potential as a prognostic biomarker. This correlation, consistent with recent studies (Warli et al., 2022; Yudiana, 2023), suggests that PD-L1 expression could be a valuable tool in risk stratification for PSCC patients. The link between PD-L1 positivity and advanced disease stages, as well as poorer clinical outcomes, highlights the urgent need for targeted therapies that can modulate the PD-1/PD-L1 axis in PSCC.

Our machine learning analysis provides a novel perspective on the factors influencing PD-L1 expression. The revelation that histologic subtype has the highest explanatory power for PD-L1 expression in tumor cells, followed by host response and histologic grade, expands on previous observations. This hierarchical influence of pathological features on PD-L1 expression points to a complex interplay between tumor biology and the immune microenvironment, which could have significant implications for immunotherapy strategies.

The role of the tumor microenvironment in modulating PD-L1 expression opens up new avenues for research. Recent studies indicating that factors such as inflammation and immune cell presence can influence PD-L1 levels (F. Li et al., 2021) suggest that targeting the tumor microenvironment could be a promising approach to enhance the efficacy of immunotherapies in PSCC.

Our analysis of CD8 expression in intratumoral and peritumoral lymphocytes reveals a nuanced picture of the immune response in PSCC. The significant associations with histologic subtypes, grades, and host inflammatory response indicates that CD8+ T cell infiltration is not a simple binary indicator but rather a complex marker influenced by multiple tumor characteristics. The observation of higher CD8 expression in certain high-grade tumors, particularly basaloid and sarcomatoid carcinomas, presents an intriguing paradox. While these tumors are generally more aggressive, the increased presence of CD8+ T cells suggests an active immune response that could potentially be harnessed for therapeutic benefit. This finding aligns with recent studies suggesting that a high density of CD8+ T cells correlates with improved clinical outcomes in PSCC (Ottenhof et al., 2018; Vries et al., 2023).

The strong association between CD8 expression and host response, as demonstrated by our machine learning model, underscores the critical role of the immune microenvironment in PSCC. This finding is supported by research indicating that the balance between CD8+ T cells and regulatory T cells within the tumor microenvironment can significantly influence disease progression (F. Li et al., 2021). Understanding this balance could be key to developing more effective immunotherapeutic approaches for PSCC.

The complex relationships between pathologic features, PD-L1, and CD8 expression revealed by our study have far-reaching implications for PSCC management. As recent studies have shown (Su et al., 2020; Warli et al., 2022), PD-L1 expression has significant potential as a predictive biomarker for immunotherapy eligibility. The association between PD-L1 expression and specific histologic subtypes and grades could guide patient selection for immunotherapeutic interventions, potentially improving response rates and patient outcomes.

Our findings on CD8 expression provide a strong rationale for exploring strategies to enhance CD8+ T cell function in PSCC. The observed correlation between CD8+ T cell infiltration and host inflammatory response suggests that modulating the tumor microenvironment could be a promising therapeutic approach. This could involve combination therapies that not only target the PD-1/PD-L1 axis but also enhance T cell recruitment and activation within the tumor microenvironment, an approach supported by recent clinical trials (Vries et al., 2023).

The integration of machine learning in our analysis represents a significant advancement in PSCC research. This approach has allowed us to quantify the impact of various pathologic features on PD-L1 and CD8 expression, providing a more nuanced understanding of their relationships. Future studies could build on this approach to develop predictive models for treatment response and patient outcomes, potentially leading to more personalized treatment strategies for PSCC patients.

While our study provides valuable insights, it is important to acknowledge its limitations. The use of tissue microarrays, while efficient for large-scale analysis, may not fully capture the heterogeneity of PD-L1 and CD8 expression within tumors. Future studies using whole-section analysis could provide a more comprehensive picture of the spatial distribution and heterogeneity of these markers within PSCC tumors.

The potential influence of HPV status on PD-L1 expression, as noted in some studies (Santos, 2024), warrants further investigation. Given the known role of HPV in a subset of PSCC cases, understanding how viral status interacts with immune marker expression could provide crucial leads for patient stratification and treatment selection. Moreover, exploring the relationship between PD-L1/CD8 expression and specific genetic alterations in PSCC could provide additional insights into the biology of this disease.

In conclusion, our study demonstrates the complex interplay between pathologic features and immune markers in PSCC, highlighting the potential of PD-L1 and CD8 as prognostic and predictive biomarkers. The integration of traditional histopathology with machine learning techniques offers a powerful approach to unraveling these complexities. As we continue to advance our understanding of the immune landscape in PSCC, these findings pave the way for more personalized and effective treatment strategies. The potential for tailored immunotherapeutic approaches based on individual tumor characteristics and immune marker profiles holds promise for improving outcomes in patients with this challenging malignancy. Future clinical trials incorporating these insights could lead to significant advancements in PSCC treatment, potentially transforming the management of this rare but aggressive cancer.

## Data Availability

All data produced is available online at https://github.com/alcideschaux/PFCK-Penis-PRY/

